# Remedial dosing recommendation for delayed or missed rivaroxaban doses for patients with non-valvular atrial fibrillation based on Monte Carlo simulation

**DOI:** 10.1101/2020.08.10.20170795

**Authors:** Xiao-qin Liu, Yi-wei Yin, Chen-yu Wang, Zi-ran Li, Xiao Zhu, Zheng Jiao

**Affiliations:** Department of Pharmacy, Shanghai Chest Hospital, Shanghai Jiao Tong University, Shanghai, China; Department of pharmacy, Huashan Hospital, Fudan University, Shanghai, China; School of Pharmacy, University of Otago, Dunedin, New Zealand

**Keywords:** Anticoagulation, Rivaroxaban, non-adherence, Monte Carlo simulation, population pharmacokinetics, pharmacodynamics

## Abstract

**Background:** Rivaroxaban is a non-vitamin K oral anticoagulant used widely for stroke prevention in patients with non-valvular atrial fibrillation (NVAF). During long-term anticoagulant therapy, delayed or missed doses are common. However, a lack of practical instructions on remedial methods has created a barrier for patients to maximise the benefit of their medications. This study aimed to explore appropriate remedial dosing regimens for rivaroxaban-treated patients with NVAF.

**Methods:** Monte Carlo simulation based on a previously established rivaroxaban population pharmacokinetic/pharmacodynamic model for patients with NVAF was employed to design remedial dosing regimens. Both the European Heart Rhythm Association (EHRA) recommendations and the model were used to establish remedial dosing regimens, which were assessed considering the on-therapy range of drug concentration, factor Xa activity, and prothrombin time under various scenarios of non-adherence.

**Results:** Recommendations of EHRA guide may not be optimal. Our findings suggested that a missed dose is taken immediately when the delay is less than or equal to 6 h; a half dose is advisable when the delay exceeds 6 h but is less than 4 h before the next dose. It is recommended to skip a dose when there are less than 4 h before the next dose. Age or renal function do not significantly influence the remedial dosing regimen.

**Conclusion:** A remedial dosing regimen based on model-based Monte Carlo simulation was systematically developed for rivaroxaban-treated patients with NVAF with poor adherence to quickly restore drug concentrations to the on-therapy range and to reduce the risk of bleeding and thromboembolism.

What is known on this topic?

- Remedial recommendations for delayed or missed rivaroxaban dose have been mentioned in package inserts and guide, but lack of solid supporting evidence.
- Monte Carlo simulation based on population analysis have been proved as an appropriate method to explore the remedial dosing strategy.

What does this paper add?

- Remedial recommendations for delayed or missed rivaroxaban in different population are established based on Monte Carlo simulation.
- The choice of optimal remedial strategy is related to delay duration.
- This paper provides a more time-specific and individualized recommendations compared with previous recommendations.

## 1 Introduction

Rivaroxaban is one of the most commonly used non-vitamin K oral anticoagulants (NOACs), and was the first oral direct factor Da (FEIa) inhibitor approved for stroke prevention in patients with non-valvular atrial fibrillation (NVAF) ^1-5^. More than two-thirds of rivaroxaban doses are excreted by the kidney as metabolites or unchanged drugs ^6,7^; therefore, dose adjustment based on renal function is required in patients with moderate renal impairment (creatinine clearance, CrCl, 30-49 mL/min) ^2^ to achieve consistent efficacy and safety, when compared with the case in individuals with normal renal function ^8^.

Patients with NVAF receiving anticoagulant therapy usually require long-term therapy; however, adherence to NOACs, including rivaroxaban, decreases over time. In a large retrospective study, adherence to rivaroxaban was 68% 3 months after treatment initiation, and decreased to 50% after 12 months of treatment ^9^. Additionally, a considerable proportion of NVAF patients are elderly (> 70 years) ^10,11^. Delayed or missed doses, which represent a major form of non-adherence in clinical practice, is common in this population of elderly patients ^12,13^. As a class of drugs with fast on/fast off characteristics ^14^, non-adherence is associated with an increasing risk of thromboembolic events ^15^. For example, a 10% decrease in dabigatran adherence, another NOAC, results in a 13% increase in all-cause mortality and stroke ^16^. Therefore, appropriate remedial dosing for patients with poor adherence is necessary to minimise the occurrence of thromboembolic events. Developing an appropriate remedial regimen is essential for the prevention of overdose-related bleeding events and underdose-related thromboembolic events caused by inappropriate remedial dosing. By providing scenario-specific instructions, patients can maximise the benefit and minimise the risk associated with rivaroxaban therapy. Several resources provide recommendations for patients who experience a delayed or missed dose. The package inserts from the United States Food and Drug Administration (US FDA) states that ‘the patient should take the missed rivaroxaban dose immediately. The dose should not be doubled within the same day to make up for a missed dose’ ^2^. Similar recommendations are found in patient information leaflets ^17^ and consumer medicine information ^18^. Furthermore, the 2018 European Heart Rhythm Association (EHRA) practical guide on the use of non-vitamin K antagonist oral anticoagulant in patients with atrial fibrillation recommends that ‘for NOACs with a once-a-day dosing regimen, a delayed dose can be taken up until 12 h after the scheduled intake. After this time point, the dose should be skipped, and the next scheduled dose should be taken’ ^19^. However, these recommendations are not supported by solid evidence from clinical studies or by extrapolation from pharmacokinetic/pharmacodynamic (PK/PD) analyses. It is also uncertain whether the recommendation can be applied to patients with impaired renal function.

Although human studies are ideal for assessing proposed remedial regimen, prospective clinical research is unethical, and post-marketing adherence data are usually inadequate to explore the effects of non-adherence and to investigate appropriate remedial doses ^20^. Monte Carlo simulations based on established population PK/PD models provide a practical approach to overcome this problem and have been successfully applied to antiepileptic, antipsychotic, and immunosuppressive agents ^21^. Therefore, this study aimed to (i) assess the effect of delayed dose on the PK/PD of rivaroxaban and (ii) explore appropriate remedial dosing regimens by Monte Carlo simulation for patients with NVAF receiving rivaroxaban.

## 2 Materials and methods

### 2.1 Patients and dosing regimen

Adult patients (aged 55 or 75 years old) with various levels of creatinine clearance (CrCl, calculated by the Cockcroft-Gault formula ^22^) were simulated, including patients with normal and moderately impaired renal function (CrCl 80 and 40 mL/min, respectively). The demographic characteristics of patients with NVAF were collected from epidemiologic reports ^10,11,23^. As indicated on the US FDA label ^2^ and as recommended in the 2018 EHRA guide ^19^, patients with CrCl ≥50 mL/min received rivaroxaban 20 mg every 24 h (q24h), and those with CrCl 30-49 mL/min received 15 mg q24h. It was assumed that patients take rivaroxaban with food at the same time every day. Demographic information and simulated dosing regimens for typical patients are listed in Table 1.

**Table 1.** Demographic characteristics of simulated patients and corresponding dosing regimens.

| ID | Age | BW | LBM <sup>a</sup> | SCr | CrCl <sup>b</sup> | Dose regimen |
| --- | --- | --- | --- | --- | --- | --- |
|  | (years) | (kg) | (kg) | (mg/dL) | (mL/min) |  |
| 1 | 55 | 70 | 55 | 1.03 | 80 | 20 mg q24h |
| 2 | 55 | 70 | 55 | 2.07 | 40 | 15 mg q24h |
| 3 | 75 | 70 | 55 | 0.79 | 80 | 20 mg q24h |
| 4 | 75 | 70 | 55 | 1.58 | 40 | 15 mg q24h |

Abbreviations: LBM, lean body mass; BW, body weight; SCr, serum creatinine; CrCl, creatinine clearance.
<sup>a</sup> Calculated lean body mass (LBM) in [kg]. for males: $LBM (kg) = 1.10 \cdot (\text{weight (kg)}) - 128 \cdot$
$\left(\frac{\text{weight (kg)}}{\text{height (cm)}}\right)^2$ ; for females: $LBM (kg) = 1.07 \cdot (\text{weight (kg)}) - 148 \cdot \left(\frac{\text{weight (kg)}}{\text{height (cm)}}\right)^2$ .
<sup>b</sup> Calculated creatinine clearance by Cockcroft-Gault formula in [mL/min]. for males: $CrCl (mL/$
$min) = \frac{(140 - \text{age (years)}) \cdot (\text{weight (kg)})}{72 \cdot SCr (mg/dL)}$ ; for females: $CrCl (mL/min) = \frac{(140 - \text{age (years)}) \cdot (\text{weight (kg)})^{0.85}}{72 \cdot SCr (mg/dL)}$ .

**Table 2.**
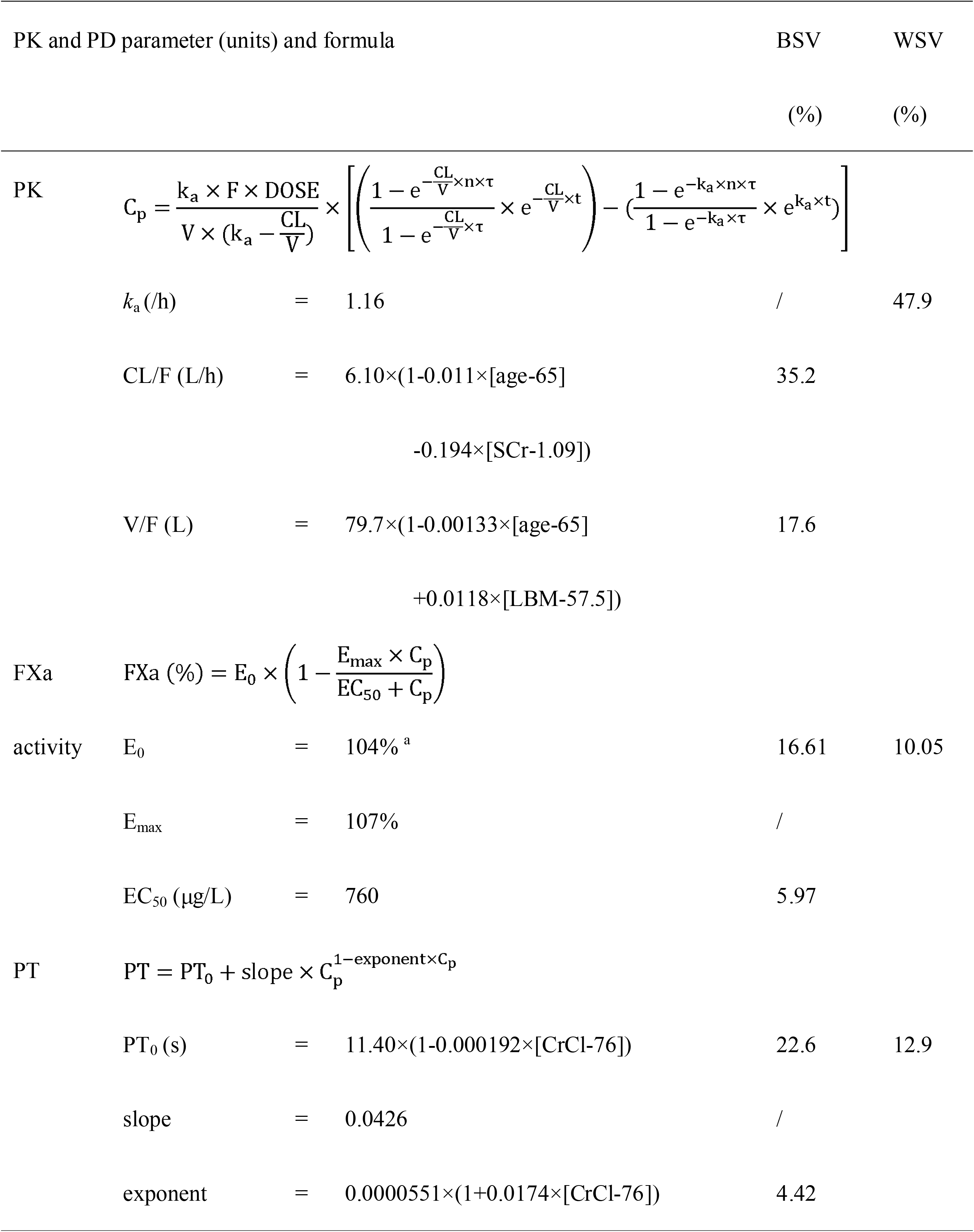
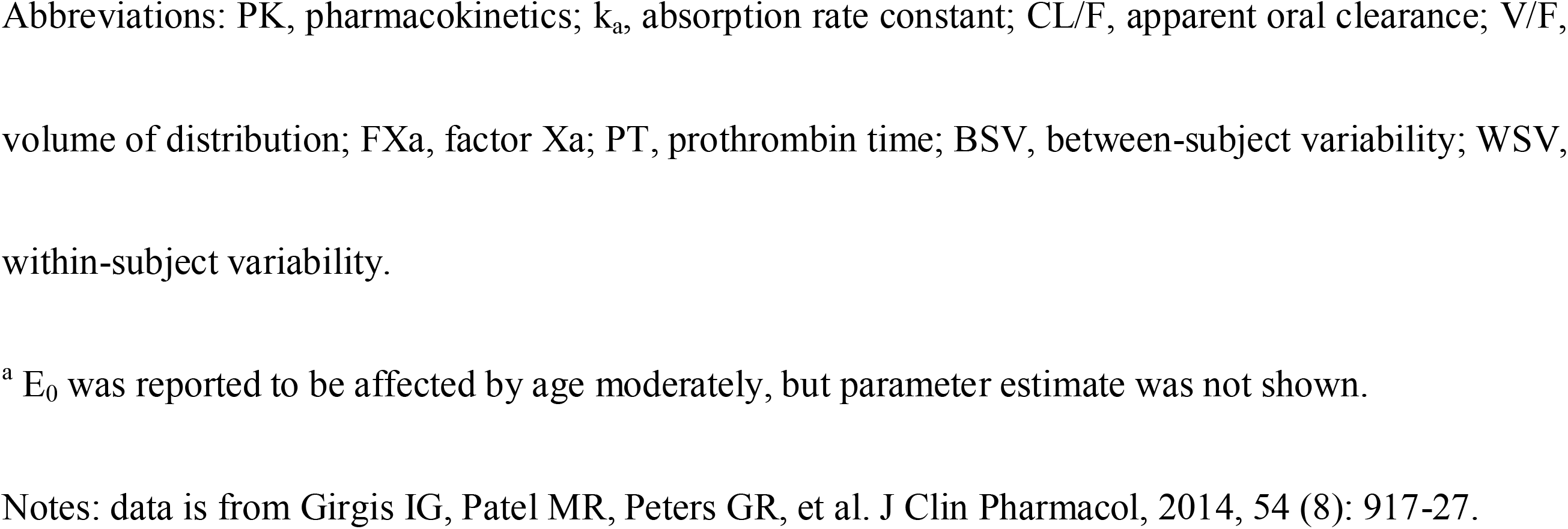
Population pharmacokinetic/pharmacodynamic (PK/PD) parameter estimates used in the Monte Carlo simulation ^24^

| PK and PD parameter (units) and formula |  | BSV | WSV |
| --- | --- | --- | --- |
|  |  | (%) | (%) |
| PK | $C_p = \frac{k_a \times F \times \text{DOSE}}{V \times (k_a - \frac{CL}{V})} \times \left[ \left( \frac{1 - e^{-\frac{CL}{V} \times n \times \tau}}{1 - e^{-\frac{CL}{V} \times \tau}} \times e^{-\frac{CL}{V} \times t} \right) - \left( \frac{1 - e^{-k_a \times n \times \tau}}{1 - e^{-k_a \times \tau}} \times e^{k_a \times t} \right) \right]$ | | |
| | $k_a$ (/h) = 1.16 | / | 47.9 |
| | $CL/F$ (L/h) = 6.10×(1-0.011×[age-65]<br>-0.194×[SCr-1.09]) | 35.2 | |
| | $V/F$ (L) = 79.7×(1-0.00133×[age-65]<br>+0.0118×[LBM-57.5]) | 17.6 | |
| FXa | $\text{FXa (\%)} = E_0 \times \left( 1 - \frac{E_{\max} \times C_p}{EC_{50} + C_p} \right)$ | | |
| activity | $E_0$ = 104% <sup>a</sup> | 16.61 | 10.05 |
| | $E_{\max}$ = 107% | / | |
| | $EC_{50}$ (μg/L) = 760 | 5.97 | |
| PT | $PT = PT_0 + \text{slope} \times C_p^{1-\text{exponent} \times C_p}$ | | |
| | $PT_0$ (s) = 11.40×(1-0.000192×[CrCl-76]) | 22.6 | 12.9 |
|  | slope = 0.0426 | / |  |
|  | exponent = 0.0000551×(1+0.0174×[CrCl-76]) | 4.42 |  |

Abbreviations: PK, pharmacokinetics; $k_a$ , absorption rate constant; CL/F, apparent oral clearance; V/F, volume of distribution; FXa, factor Xa; PT, prothrombin time; BSV, between-subject variability; WSV, within-subject variability.
<sup>a</sup> E<sub>0</sub> was reported to be affected by age moderately, but parameter estimate was not shown.
Notes: data is from Girgis IG, Patel MR, Peters GR, et al. J Clin Pharmacol, 2014, 54 (8): 917-27.

### 2.2 Remedial dosing regimen assessment

For patients with NVAF taking rivaroxaban 20 or 15 mg daily, various scenarios were investigated in which doses were delayed from 1 to 24 h after the scheduled time (Figure 1).

**Fig. 1.**
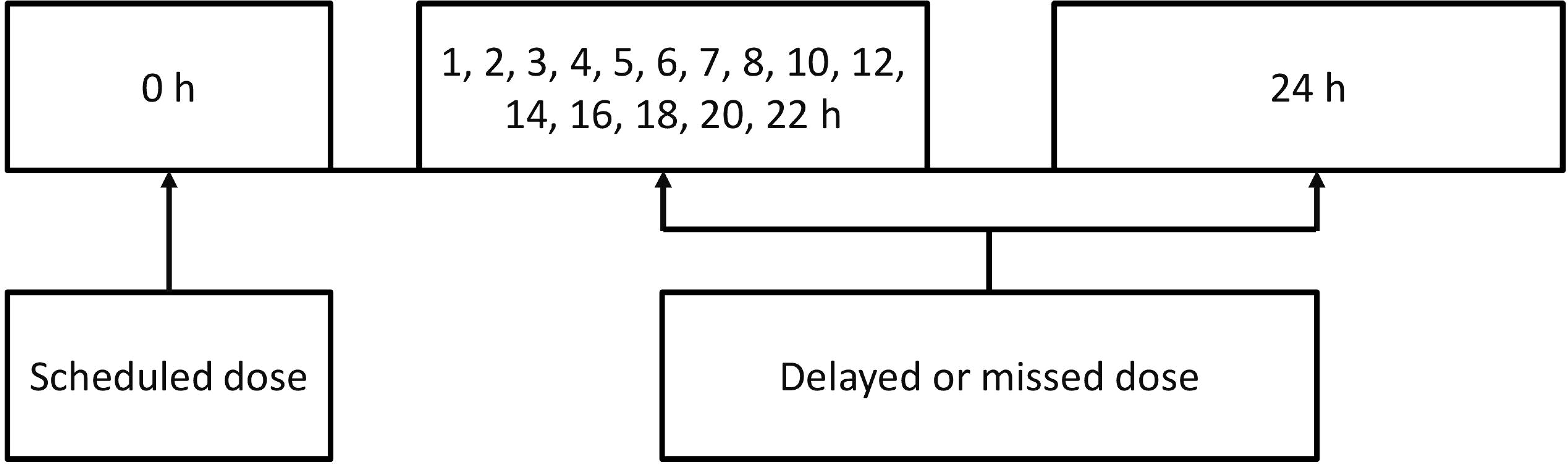
Simulation scenarios for delayed or missed rivaroxaban doses for patients with non-valvular atrial fibrillation (NVAF) receiving 15 or 20 mg dose every 24 h.

#### 2.2.1 Population PK/PD model and simulation

A previously published population PK/PD analysis of rivaroxaban was used in our study ^24^. The study cohorts were obtained from a subset of patients in the largest multinational phase III ROCKET AF trial (Rivaroxaban once daily Oral direct factor Xa inhibition Compared with vitamin K antagonism for prevention of stroke and Embolism Trial in Atrial Fibrillation) ^25^. A total of 161 patients with NVAF, with time-matched PK and PD samples were included in this population PK/PD analysis. These included 136 patients with normal or mildly impaired renal function (CrCl≥50 mL/min) receiving 20 mg daily, and 25 patients with moderate renal impairment (CrCl 30–49 mL/min) receiving 15 mg daily. Detailed demographic and dosing regimen information is listed in Supplementary Table 1. Blood samples were used to determine rivaroxaban concentration, Factor Xa (FXa) activity, and prothrombin time (PT).

In this analysis, population PK was depicted by a one-compartment model with first-order absorption and elimination. The PK model was parameterised by the absorption rate constant (k_a_), apparent clearance (CL/F), and apparent volume of distribution (V/F). The time-concentration relationship is described in Eq. 1.

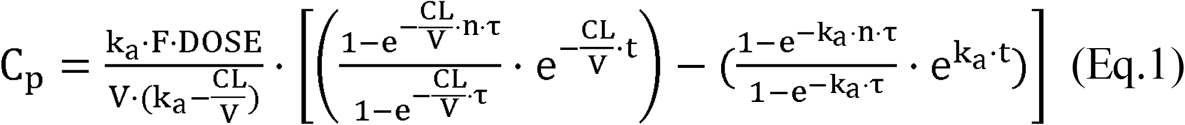

This equation comprised bioavailability (F), dose (DOSE; mg), apparent volume of distribution (V; L), number of doses administered (n), dosing interval (τ; h), time after the last doset; h), and concentration at time t(C_p_; mg/L). Age and serum creatinine (SCr; mg/dL) affected CL/F (Eq 2.), while age and lean body mass (LBM) influenced V/F (Eq 3.).

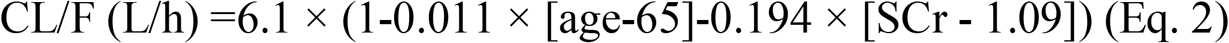

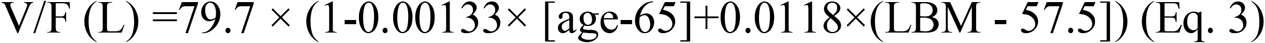

PD markers, including FXa activity and PT, were modelled to correlate with rivaroxaban concentration. FXa activity was negatively correlated with the concentration of rivaroxaban, with a direct inhibitory maximum-effect (E_max_) relationship, as described by Eq 4.

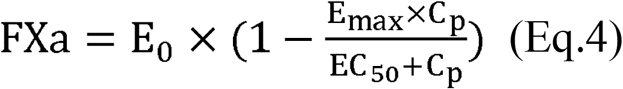

where, E_0_ represents baseline FXa activity, E_max_ represents the relative maximum level of inhibition, and EC_50_ represents the concentration of rivaroxaban resulting in 50% of maximum inhibition.

PT was positively correlated with rivaroxaban concentration in a near-linear relationship, as described by Eq 5.

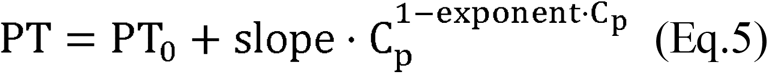

PT_0_ represents baseline PT, slope represents the per unit difference in CrCl (mL/min) to the median CrCl (76 mL/min), and the exponent represents the parameter describing the decline linearly with increasing C_p_. CrCl was found to affect both PT_0_ and the exponent, which is expressed by Eq.6 and Eq.7.

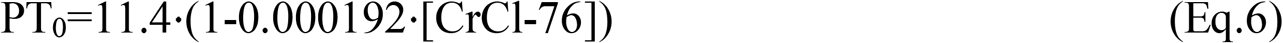

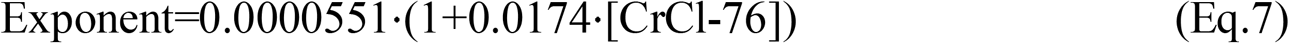

All PK/PD parameter estimates employed in the Monte Cario simuiation are iisted in Tabie 2.

Monte Cario simuiations were performed with NONMEM software (version 7.4; Icon Incorporation, PA, USA) using the $SIMULATION biock. Output profiies were processed using the R package (version 3.5.3, https://www.r-project.org/). A totai of 5000 virtuai patients were simuiated to depict the PK/PD profiles under each non-adherence scenario. Virtual patients were assumed to have aiready received muitipie doses of rivaroxaban and have reached steady state, and to obtain the expected ievei of anticoaguiation without any unexpected drug-related adverse effects.

#### 2.2.2 Remedial dosing regimens

Based on previous research and on ciinicai practise ^21^, six strategies, as weii as recommendations inciuded in the EHRA guide, assessed when the dose was deiayed or missed. Graphic remedial strategies are shown in Figure 2, and details are below as follows:

Strategy A: skip the delayed dose, administer the regular dose at the next scheduled dosing time, and then resume the regular dosing regimen.
Strategy B: administer the regular dose immediately, followed by a regular dose at the next scheduled dosing time, and then resume the regular dosing regimen.
Strategy C: administer a half dose immediately followed by a regular dose at the next scheduled dosing time, and then resume the regular dosing regimen.
Strategy D: administer a regular dose immediately followed by a half dose at the next scheduled dosing time, and then resume the regular dosing regimen.
Strategy E: administer one and a half regular doses immediately, skip the next scheduled dose, and then resume the regular dosing regimen.
Strategy F: administer a double dose, skip a dose at the next scheduled dosing time, and then resume the regular dosing regimen;
EHRA guide: administer the delayed dose if less than 12 h late; otherwise, skip the delayed or missed dose and administer the next scheduled dose.

**Fig. 2.**
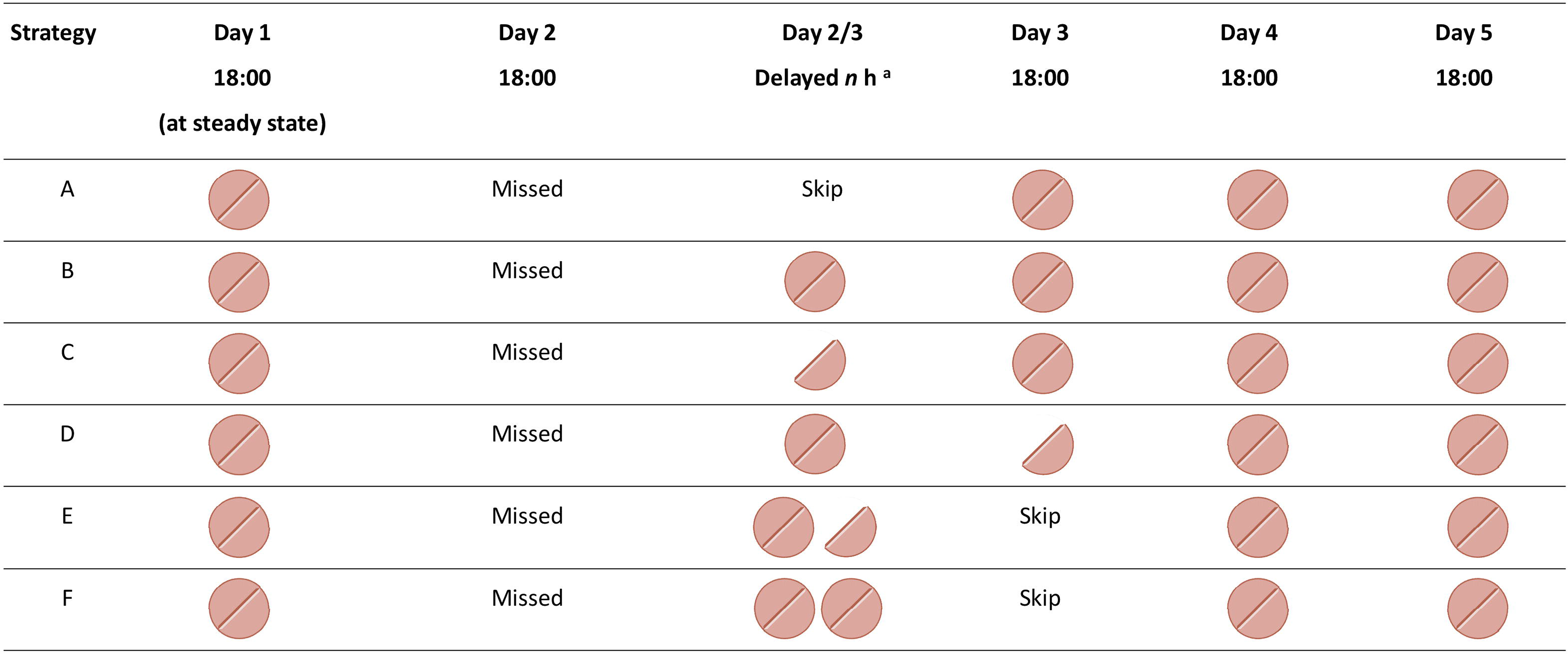
Graphical representation of six remedial dosing regimens following a delayed or missed dose. 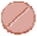 Represents an integrated rivaroxaban dose, and 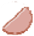 represents a half rivaroxaban dose. Patients were assumed to take rivaroxaban at 18:00 daily and to have reached a steady state following a delayed or missed dose. *^a^n* represents the delayed time, whose range is from 1 to 24 h.

#### 2.2.3 Index for evaluating remedial regimens

Currently, there is no widely accepted therapeutic range of rivaroxaban concentrations, FXa activity, or PT. Therefore, the on-therapy range, defined as the ‘interval delineated by the 5^th^ percentile trough concentration and the 95^th^ percentile peak concentration’ at the steady state of a given dose ^26,27^, was used in this study. Considering the differences in patient demographics, we estimated the on-therapy range for each typical patient at steady state by Monte Carlo simulation. To assess the effect of a delayed or missed dose, we defined the percentage of individuals outside the on-therapy range as those whose concentration or PD marker levels were not within the on-therapy range. The percentages of individuals outside the on-therapy range under all non-adherence scenarios described above were estimated to evaluate the effect of non-adherence over time.

Deviation time, defined as the duration when simulated rivaroxaban concentration, FXa activity or PT are outside the on-therapy range, was chosen as the index to evaluate the remedial regimen. It was calculated by adding each deviation time when simulated data is higher than the upper limit and lower than the lower limit of on-therapy range. The two aspects of deviation time may represent higher risk of bleeding and thromboembolic events, respectively. It was assumed that patients taking regular rivaroxaban doses had reached expected anticoagulant effects within the on-therapy range. Therefore, when the rivaroxaban dose was delayed or missed, optimal remedial dosing could help patients to restore the rivaroxaban concentration, FXa, or PT to the on-therapy range as soon as possible to minimise the deviation time. Two remedial dosing regimens with deviation times of less than 1 h were assumed to be equivalent. If there were discrepancies among recommendations for rivaroxaban concentration, FXa activity, or PT, those based on FXa activity were prioritised. This was because quantification of FXa activity is considered as a more specific biomarker for NOACs, which is recommended by the International Society on Thrombosis and Haemostasis American College of Chest Physicians guidelines ^28,29^.

#### 2.2.4 Sensitivity analysis

A sensitivity analysis was applied to identify parameters that had a substantial influence on model output, and to determine the extent that these important parameters contributed to the overall variability in model output ^30^. Therefore, dosing intervals and demographic characteristics (age and body weight) were included in the sensitivity analysis to investigate their influence on the remedial dosing regimens.

Irregular dosing intervals of 23–25 h (18:00, 17:00, and 18:00) and 25–23 h (18:00, 19:00, and 18:00) were assessed considering situations where patients took rivaroxaban at different times every day. Moreover, considering the demographics of the modelling population, patients with different ages and body weights (expressed as LBM in population analysis) were also investigated (Supplementary Table 2). Furthermore, the criterion used to judge the equality of the two strategies was also examined from 0.5 to 2 h, to investigate whether it had an influence on the remedial dosing recommendations.

## 3 Results

### 3.1 Effect of delayed or missed doses of rivaroxaban on PK/PD

The results from the Monte Carlo simulation showed that the effect of a delayed dose was related to the delay time. With an increasing delay time, the risk of a patient being outside the on-therapy range was also increased considering both the PK and PD (Figure 3). For example, among patients with CrCl of 80 mL/min who took rivaroxaban 20 mg q24h, 12.6% and 47.9% patients were outside the on-therapy range based on rivaroxaban concentration when the dose was delayed for 6 h and 24 h, respectively.

**Fig. 3.**
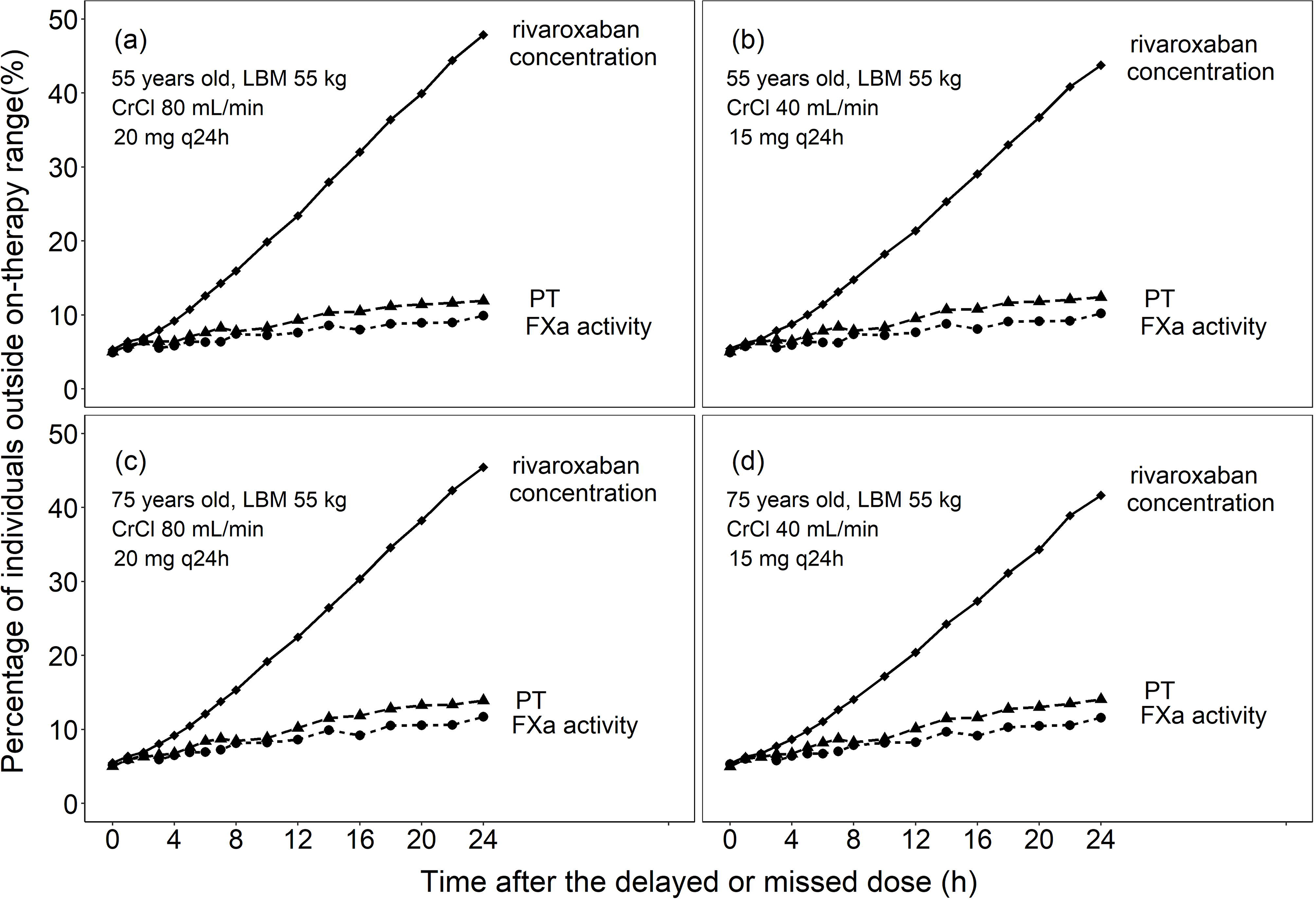
Percentage of patients with NVAF falling outside the on-therapy ranges *versus* time after the delayed or missed doses in terms of rivaroxaban concentration, FXa activity and PT. (a) Adult patients, 55 years old, CrCl 80 mL/min receiving 20 mg every 24 h (q24h). (b) Adult patients, 55 years old, CrCl 40 mL/min receiving 15 mg q24h. (c) Elderly patients, 75 years old, CrCl 80 mL/min receiving 20 mg q24h. (d) Elderly patients, 75 years old, CrCl 40 mL/min receiving 15 mg q24h. Simulated patients were assumed to take rivaroxaban doses regularly and to have reached steady state.

The effect of delayed dose was not significantly affected by age or renal function. (Figure 3). For example, among elderly patients with normal renal function and impaired renal function, the percentage of those who were outside the on-therapy concentration range was 22.6% and 20.4% when the dose was delayed for 12 h, respectively, representing a difference less than 15%. Moreover, the difference in the percentages of patients who were outside of the FXa activity (8.6 vs. 8.26%) or PT on-therapy (10.2 vs. 10.1%) range were also less than 15%.

In addition, the impact of the delayed dose on the PK/PD was very similar when the delay time was less than 3 h. However, the difference increased as the delay time increased (Figure 3). Considering rivaroxaban concentration, this percentage increased approximately linearly with time. The percentage changes considering FXa activity and PT were close, with a smaller slope and increased with the delay time. For example, for adult patients receiving a 20 mg q24h dose delayed for 2 h, the percentages of those outside the on-therapy range were 6.9, 6.5, and 6.4% for rivaroxaban concentration, FXa activity, and PT, respectively. These percentages were 23.4, 7.6, and 9.3%, respectively, when dosing was delayed for 12 h, with a greater difference between rivaroxaban concentration and PD markers compared with that at 2 h.

### 3.2 Remedial dosing regimens

The remedial dosing recommendations following a delayed rivaroxaban dose are summarised in Table 3. Figure 4a–c shows the concentration, FXa activity, and PT profiles for fully adherent elderly patients with impaired renal function (aged 75 years, CrCl 40 mL/min, receiving 15 mg q24h). In general, remedial dosing recommendations are related to delayed time. A regular dose of rivaroxaban could be taken immediately when the delay doesn’t exceed 6 h. When the delay exceeds 6 h but is less than 20 h, it is advisable to remedy a half dose either by taking a half dose immediately followed by a regular dose at the next scheduled time (6-14 h), or by taking a regular dose immediately followed by a half dose at the next scheduled dosing time (14–20 h). When the delay exceeds 20 h, it is recommended to skip the delayed dose and take a regular dose at the next scheduled dosing time. When a dose is missed, it is advisable to take a regular dose at the scheduled dosing time.

**Table 3.**
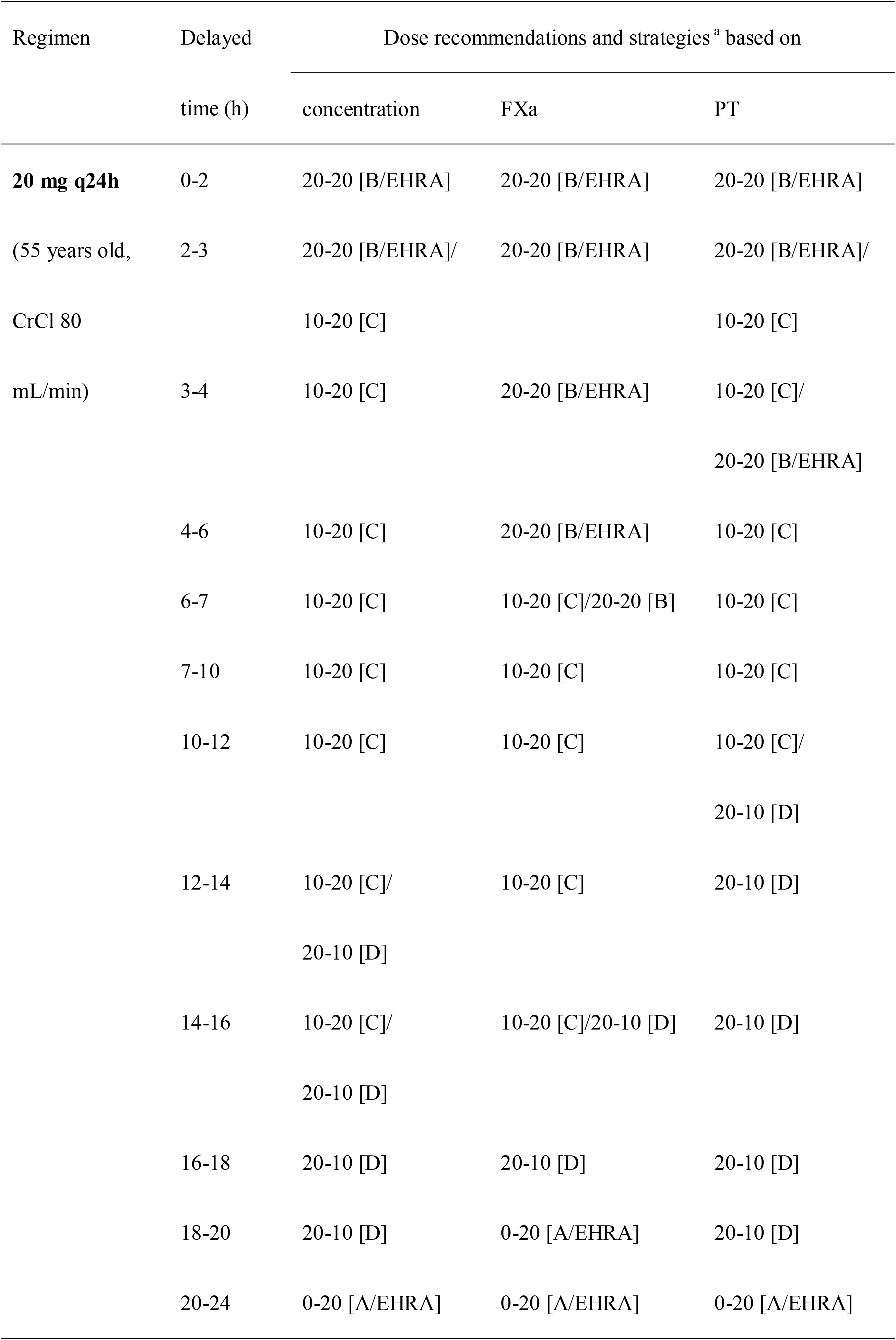
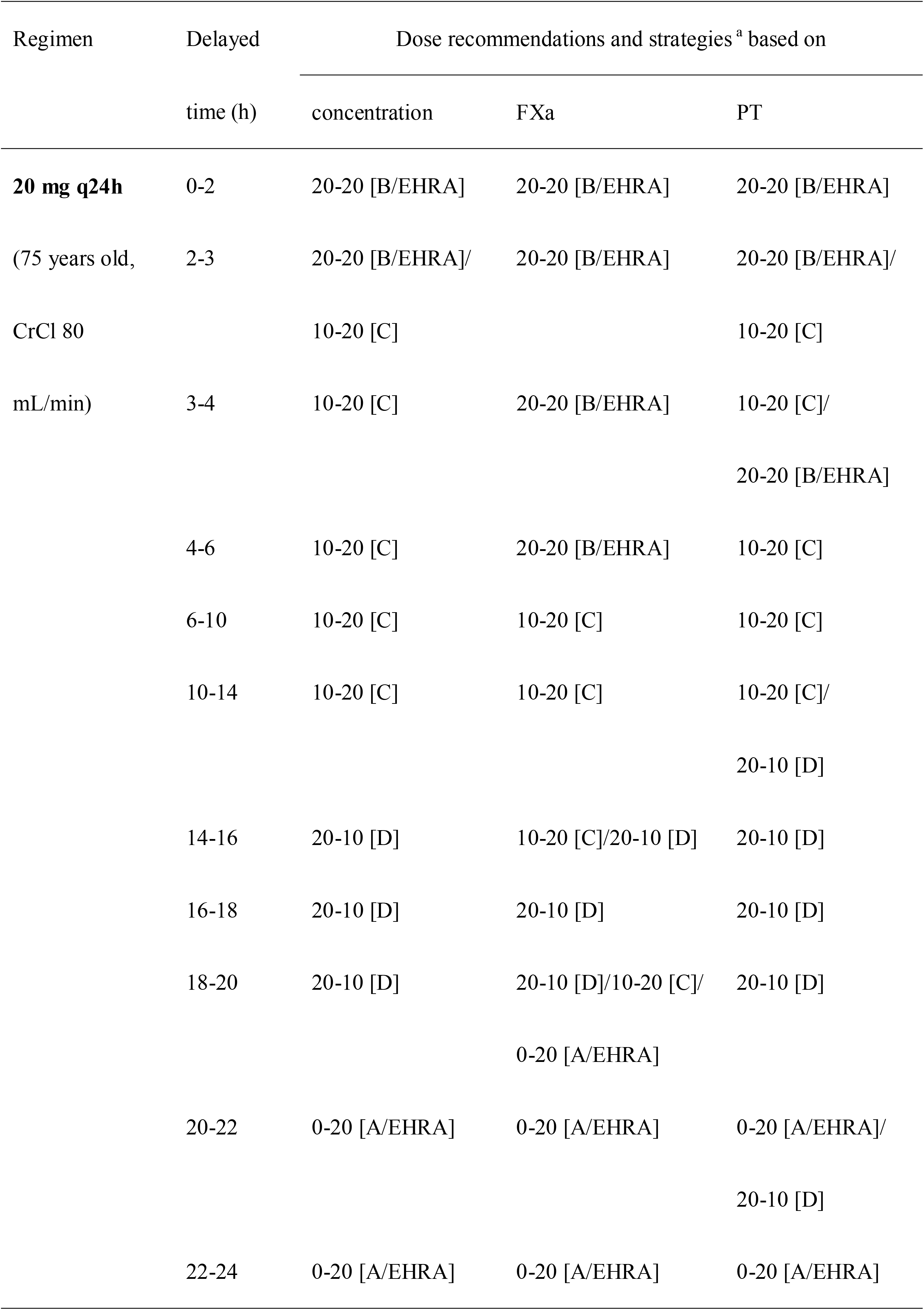
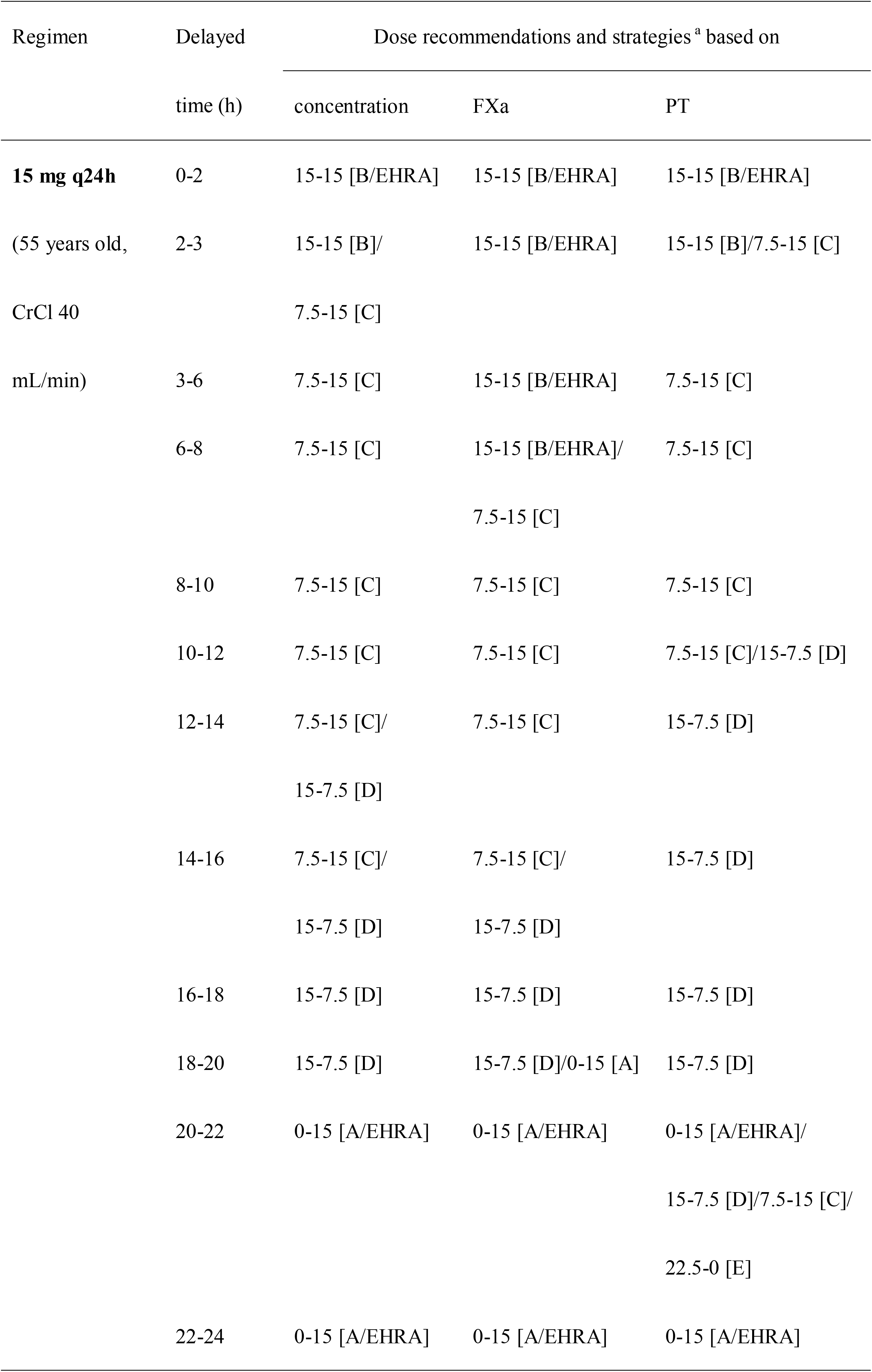
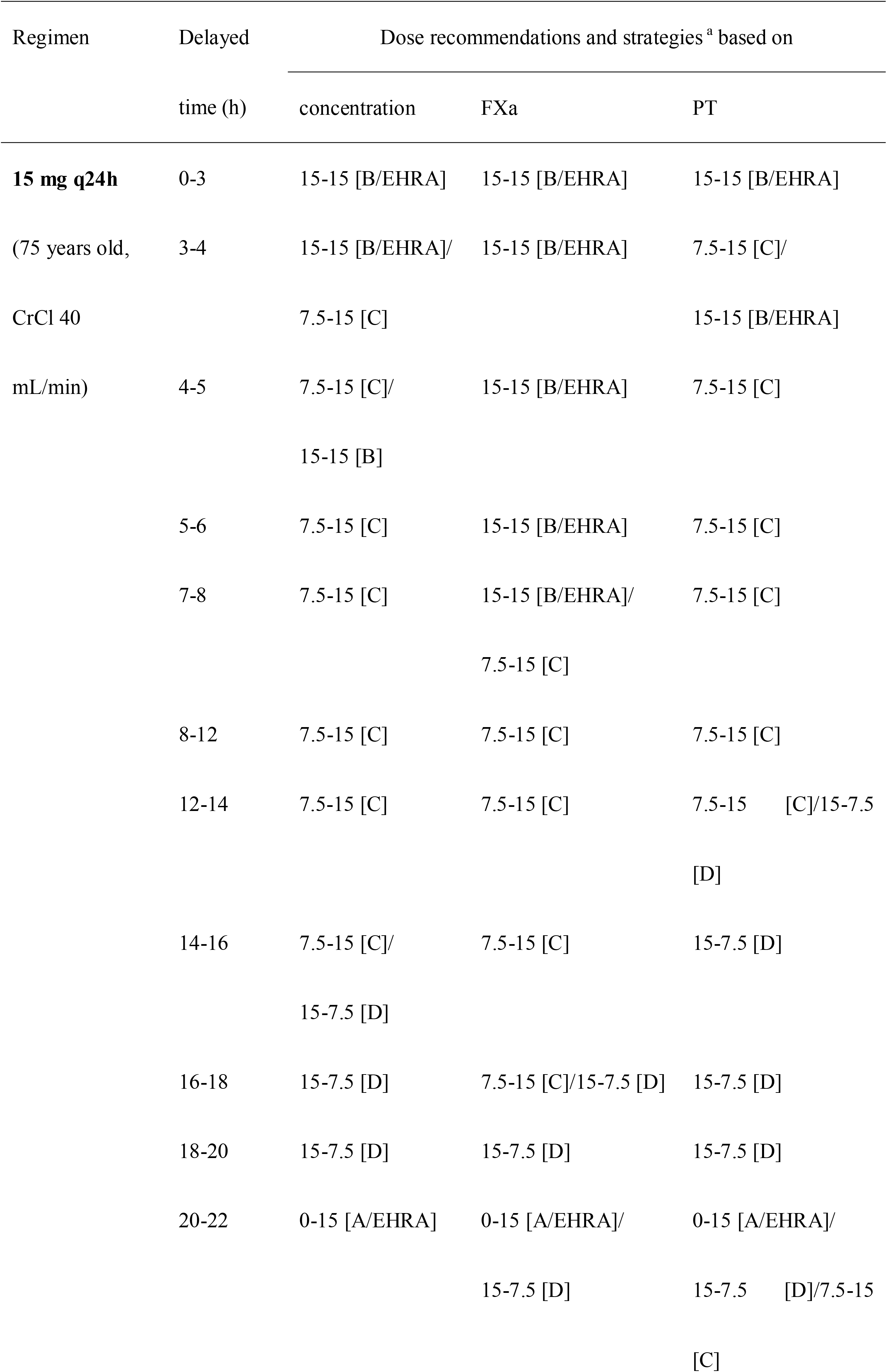
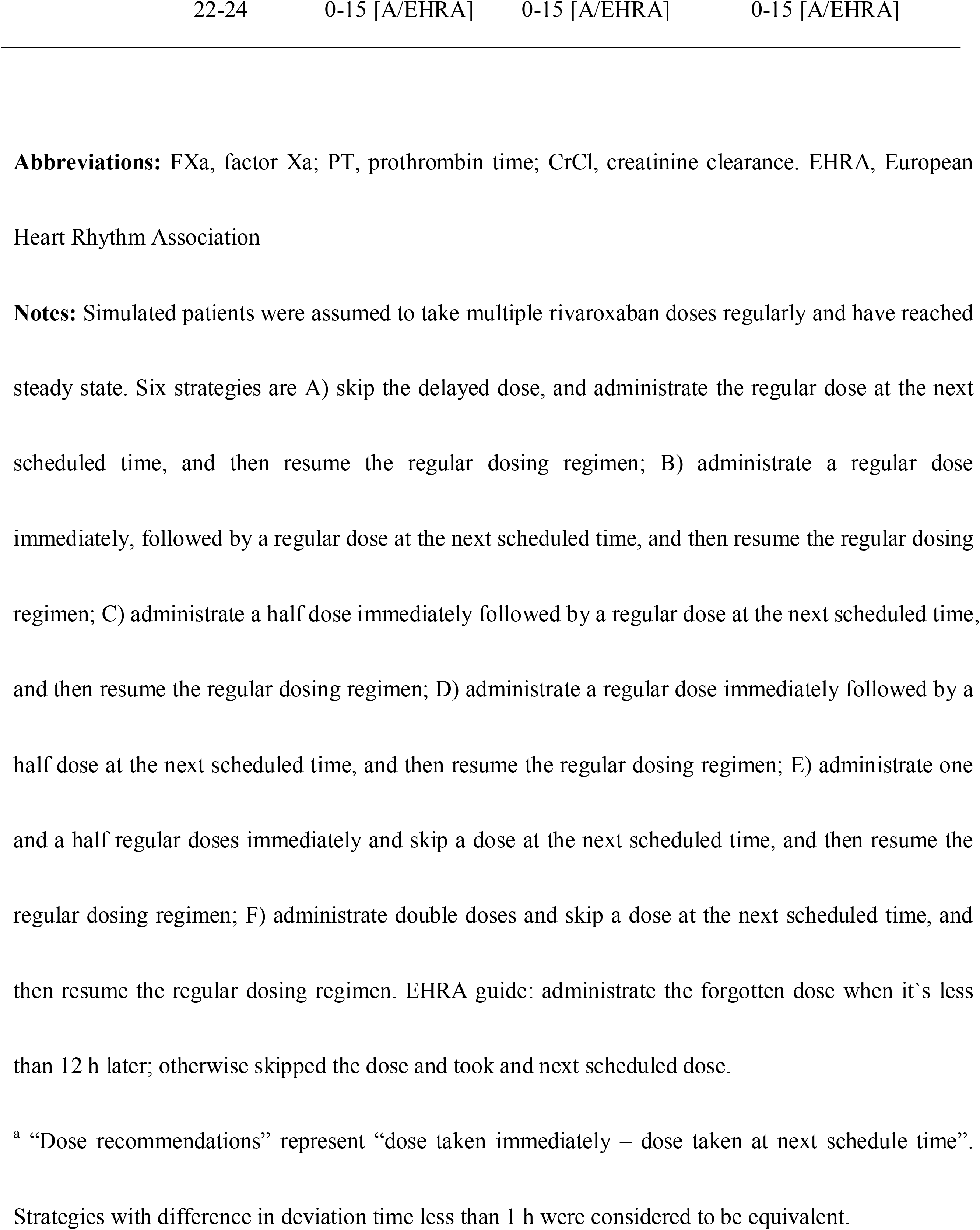
Remedial dosing recommendations for patients with atrial fibrillation

**Fig. 4.**
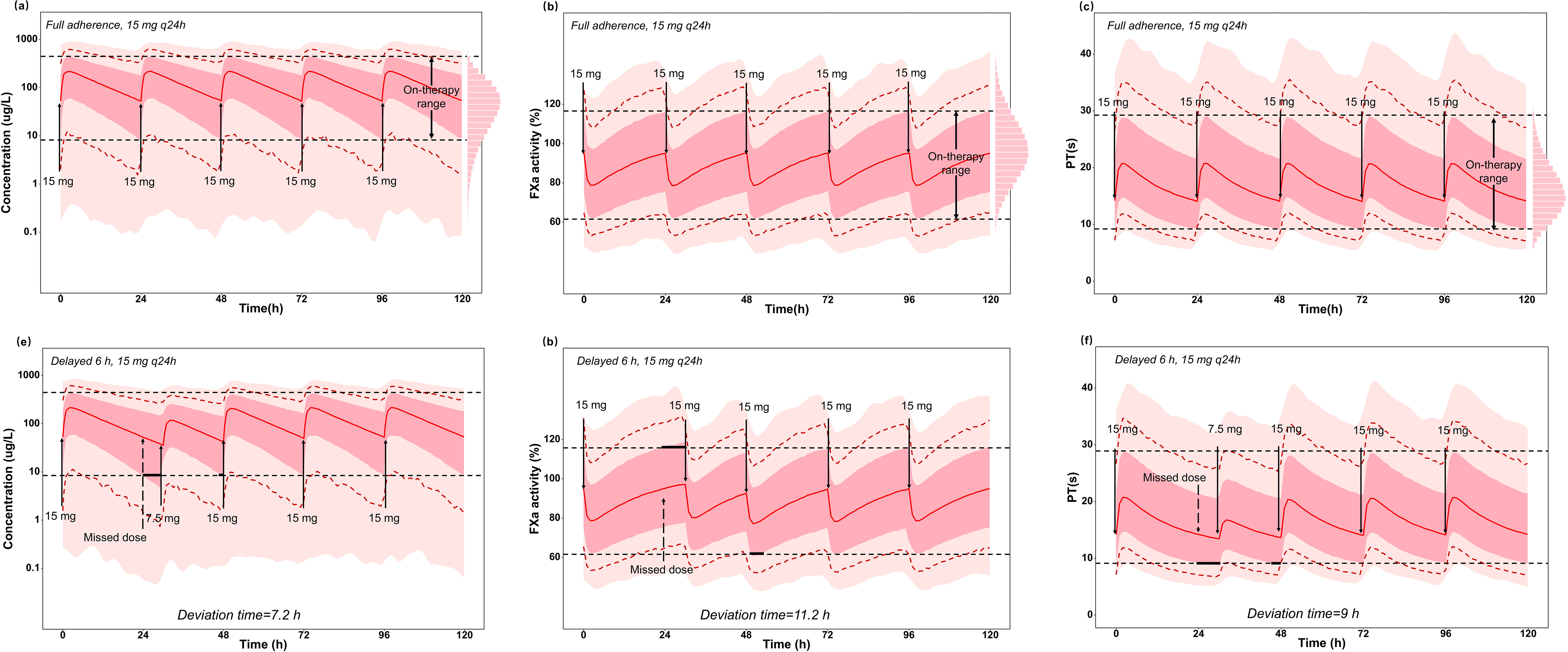
Pharmacokinetic (PK) and pharmacodynamic (PD) profiles under full adherence and optimal remedial regimens in terms of rivaroxaban concentration, FXa 30 activity and PT for elderly patient with CrCl 40 mL/min receiving rivaroxaban 15 mg every 24 h when the dose was delayed for 6 h. (a) Full adherence based on concentration, (b) FXa activity, and (c) PT. (d) optimal remedial regimens in terms of concentration, (e) FXa activity, and (f) PT when the dose was delayed for 6 h. Dark pink shadows represent the distribution of the range between the 5th percentile of the simulated trough concentration (FXa activity or PT) and 95th percentile of the simulated peak concentration (FXa activity or PT). Light pink shadows represent the distribution of the remaining simulated concentration (FXa activity or PT). Red solid lines represent the median of the simulated concentration (FXa activity or PT). The red dotted lines represent the 0.5th percentile and 99.5th percentile of the simulated concentration (FXa activity or PT). Black dotted lines represent the on-therapy range calculated by the simulated concentration (FXa activity or PT). Black horizontal bold solid lines represent the deviation time.

Recommendations based on rivaroxaban concentration and FXa or PT levels were the same among almost all scenarios, except for scenarios where the delay time was 3–6 h. Rivaroxaban concentration, FXa activity, and PT profiles following a delay of 6 h under optimal remedial regimens for elderly patients with impaired renal function (aged 75 years, CrCl 40 mL/min, taking 15 mg q24h) are shown in Figure 4d–f. Under this scenario, recommendations based on FXa activity supported taking a regular dose immediately, while recommendations based on concentration and PT preferred taking a half dose. Based on pre-established criteria where FXa activity was preferred, recommendations based on FXa activity were selected.

Additionally, age and renal function did not affect remedial dosing regimens. For example, for adult patients with normal renal function, a half dose (10 mg) is recommended when the dose is delayed by 6–20 h, based on rivaroxaban concentration, FXa activity, and PT. For elderly patients with impaired renal function taking a lower daily dose, a half dose (7.5 mg) is also recommended when the delay was 6–20 h.

Because recommendations from the EHRA did not consider splitting rivaroxaban tablets, this represented a missed opportunity to provide a more personalised remedial strategy. Based on the present results, our recommendation is consistent with that of the EHRA only when the delay was less than 3 h, or it was less than 4 h before the next dose. Recommendations from the EHRA guide are not optimised for most scenarios. A comparison of the deviation time between the EHRA guide and our proposed remedial strategy is shown in Figure 5. Deviation times under different strategies are listed in Supplementary Table 3–6, which can provide supportive information to balance the risk of thromboembolism and bleeding.

**Fig. 5.**
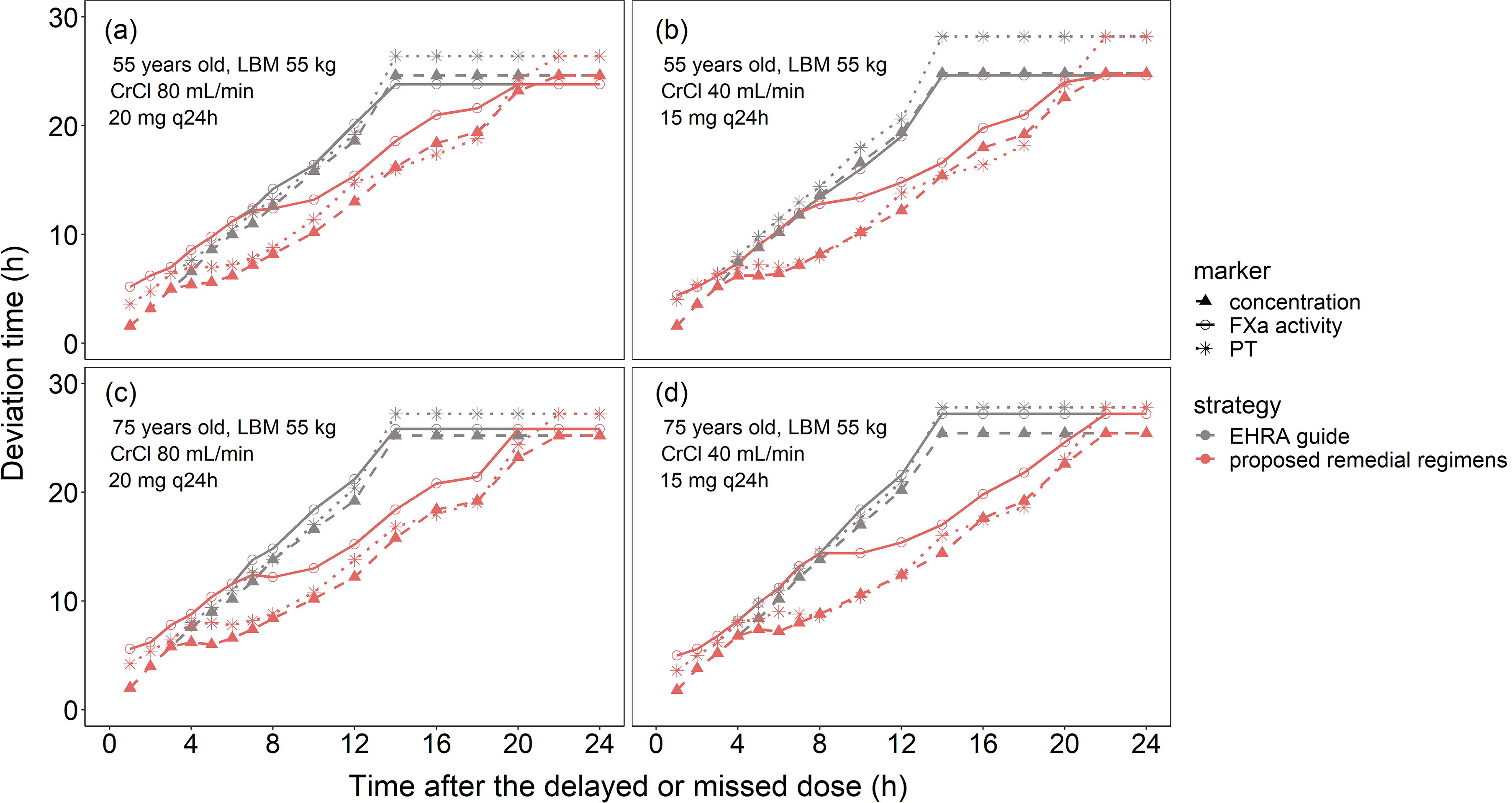
Total deviation time from recommendations by the European Heart Rhythm Association (EHRA) guide and proposed remedial regimens. (a) Adult patients, 55 years old, CrCl 80 mL/min taking 20 mg every 24 h (q24h). (b) Adult patients, 55 years old, CrCl 40 mL/min taking 15 mg q24h. (c) Elderly patients, 75 years old, CrCl 80 mL/min taking 20 mg q24h. (d) Elderly patients, 75 years old, CrCl 40 mL/min taking 15 mg q24h. Simulated patients were assumed to take rivaroxaban doses regularly and to have reached steady state.

### 3.3 Sensitivity analysis

Age, body weight, dosing intervals, and criterion for assessing the equality of the two strategies were included in the sensitivity analysis when the rivaroxaban dose was delayed for 3, 6, 12, and 24 h. Deviation time may vary slightly by age, body weight, and dosing intervals. However, these factors did not significantly impact remedial dosing regimens (Supplementary Figure 1). Broadening the criterion for assessing the equality of the two strategies resulted in more options for remedial strategies at some time points, whereas narrowing the criterion resulted in fewer options (Supplementary Tables 7–10). As shown in Table 3, for adult patients with normal renal function, recommendations included 10 mg with strategy C or 20 mg with strategy B when there was a delay of 4 h considering the PT. However, the recommended strategy only included 10 mg with strategy C when the criteria were narrowed. When the criteria were broadened, 10 mg with strategy D was included in the recommended strategies.

## 4 Discussion

This was the first study to characterise the effects of delayed rivaroxaban dose on the PK/PD profiles and to investigate appropriate remedial dosing regimens for patients with NVAF under different scenarios. A few studies have estimated the rate of non-adherence of NOACs and explored the associated risk factors ^12,13,15^. However, non-adherence is common, and very few studies have proposed solutions when doses are delayed or missed. Considering the high risk for thromboembolism resulting from non-adherence and the serious consequences resulting from inappropriate remedial and missing/delayed doses, our study is the first to provide insight into remedial regimens for patients with NVAF following delayed or missed rivaroxaban doses.

Our study explored optimal remedial regimens based on Monte Carlo simulation from the perspective of both PK and PD, and considering both population level and individual variation. Handling delayed doses from the population PK perspective has been previously discussed for various diseases, such as epilepsy^31-34^, schizophrenia ^35,36^ and renal transplantation ^37^. However, few studies have explored this with consideration of PD, or PK/PD ^21^. The PD effect should also be considered for medicine acting via an indirect mechanism ^38^, such as rivaroxaban, which acts as an indirect anticoagulant by inhibiting FXa activity.

Multiple PK and PD markers were used in our study, including rivaroxaban concentration, FXa activity, and PT. Given that no clearly defined therapeutic ranges have been established for rivaroxaban ^26^, an on-therapy range was adopted to describe appropriate maintained levels of concentration, FXa activity, and PT. The concept of an on-therapy range has been used for antiepileptic drugs, and was thought to be appropriate for epilepsy management ^32,33^. Regarding rivaroxaban, the calculated PK on-therapy ranges based on simulated data are close to those estimated in clinical trials ^39^. We hypothesised that patients taking the prescribed dose could obtain expected anticoagulation outcomes within the on-therapy range. It is reasonable to assume that a remedial strategy restoring drug concentrations to a previous range can minimise negative outcomes associated with non-adherence, such 0061s lower exposure, in addition to potential thromboembolic events.

Through population analysis and Monte Carlo simulation, our study provides remedial dosing recommendations that are more time-specific than those included in the EHRA guide. Recommendations from the EHRA are general for all NOACs, including rivaroxaban, apixaban, edoxaban, and dabigatran ^19^. Our research optimises these recommendations from both PK and PD perspectives, suggesting that the cut-off point for remedying a full dose is not 12 h, as noted in the EHRA guide. Based on the concentration and PT, the cut-off point is approximately 3 h, while based on FXa activity, the cut-off is approximately 6 h. When the dose is delayed for less than the cut-off point, taking the delayed dose of rivaroxaban as soon as possible is recommended. However, when the delay exceeds the cut-off point, a half dose is advisable, to minimise the deviation time. When the dose is close to the next scheduled dose or is completely skipped, it is advisable to skip the missed dose and to continue with the regular dosing regimen at the next scheduled time.

It is also notable that doubling the dose of rivaroxaban after a missed dose (strategy F) is not recommended based on the drug package insert, the EHRA guide, and our research. In fact, even remedying a half dose (strategy E) is unnecessary. This is inconsistent with the findings of previous similar studies exploring remedial dosing strategies in simulations based on antiepileptic drugs ^32-34^ or immunosuppressants ^37^, which are taken every 12 or 24 h. Remedying no extra dose when missing a dose may result in a deviation time that is below the lower limit of the on-therapy range, while remedying a half dose increases the deviation time above the upper limit of the on-therapy range. Furthermore, a double dose substantially increases the drug concentration to levels beyond the upper limit of the on-therapy range. This also leads to a much longer deviation time outside the on-therapy range.

The risk of bleeding or thromboembolism varies among patients with atrial fibrillation. The CHA2DS2-VASc (congestive heart failure, hypertension, age ≥ 75 years, diabetes mellitus, stroke or transient ischaemic attack [TIA], vascular disease, age 65–74 years, sex category) score is commonly used for the assessment of stroke risk ^40^. The risk of stroke may differ for patients of the same age and with the same renal function, considering other factors included in the CHA2DS2-VASc scoring system. Adherence to anticoagulant therapy, such as with rivaroxaban, is particularly important for patients with a high CHA2DS2-VASc score, given their high risk for stroke ^38^. For example, in patients with a history of bleeding, it is important to avoid an inappropriate remedial strategy. Therefore, deviation times that exceed the upper limit of the on-therapy range and that were below the lower limit of the on-therapy range, representing the risk of bleeding and thromboembolism, respectively, are also listed in Supplementary Tables 3–6. Therefore, clinicians can select the optimal remedial dose for individual patients based on deviation time and patient characteristics.

There are several limitations to this study. First, the simulated dosing regimen complied with that of the modelling population, who took 20 or 15 mg daily rivaroxaban based on renal function. Therefore, our recommendations may not apply to patients receiving other doses of rivaroxaban. Second, our recommendations were based on a 90% predicted interval of simulation data. Therefore, attention should be paid to special populations, such extremely obese patients or older patients, who were not included in our analysis.

In conclusion, this study assessed the effect of non-adherence to rivaroxaban and explored appropriate remedial dosing regimens for patients with NVAF considering the drug concentration, FXa, and PT. According to our model-informed analysis, the optimal remedial strategy is only dependent on the duration of the delay. We recommend that a rivaroxaban dose is taken following a delay of no more than 6 h, while a half dose may be recommended after 6 h up to a maximum of 4 h before the next dose. No dose is advisable to remedy a missed dose. Clinicians should also balance the risk of bleeding and stroke and select appropriate remedial strategies based on the clinical situation of each patient.

## Data Availability

Data of parameter estimates for simulation is from Girgis IG, Patel MR, Peters GR, et al. J Clin Pharmacol, 2014, 54 (8): 917-27.

https://accp1.onlinelibrary.wiley.com/doi/pdf/10.1002/jcph.288

## Acknowledges

We thank Dr. I.G. Girgis from Janssen Pharmaceuticals Research & Development, Raritan, USA for providing details and active discussions on the coding. We thank Hai-ni Wen MPharm., and Yun-peng Guo MPharm. from Shanghai Chest Hospital for their critical comments. We would also thank Editage (www.editage.cn) for English language editing.

## Author contributions

Xiao-qin Liu, Yi-wei Yin, and Zheng Jiao designed the article and planned the work for the manuscript. Xiao-qin Liu, Chen-yu Wang, Zi-ran Li performed the data analysis. Xiao-qin Liu, Yi-wei Yin, Xiao Zhu, and Zheng Jiao drafted and revised the manuscript. All authors approved the finial version of this manuscript.

